# Clinical utility of Elecsys Anti-SARS-CoV-2 S assay in COVID-19 vaccination: An exploratory analysis of the mRNA-1273 phase 1 trial

**DOI:** 10.1101/2021.10.04.21264521

**Authors:** Simon Jochum, Imke Kirste, Sayuri Hortsch, Veit Peter Grunert, Holly Legault, Udo Eichenlaub, Basel Kashlan, Rolando Pajon

## Abstract

**Background:** The ability to quantify an immune response after vaccination against severe acute respiratory syndrome coronavirus 2 (SARS-CoV-2) is essential. This study assessed the clinical utility of the quantitative Roche Elecsys^®^ Anti-SARS-CoV-2 S assay (ACOV2S) using samples from the 2019-nCoV vaccine (mRNA-1273) phase 1 trial (NCT04283461).

**Methods:** Samples from 30 healthy participants, aged 18–55 years, who received two injections with mRNA-1273 at a dose of 25 μg (n=15) or 100 μg (n=15), were collected at Days 1 (first vaccination), 15, 29 (second vaccination), 43 and 57. ACOV2S results (shown in U/mL – equivalent to BAU/mL per the first WHO international standard) were compared with results from ELISAs specific to antibodies against the Spike protein (S-2P) and the receptor binding domain (RBD) as well as neutralization tests including nanoluciferase (nLUC_80_), live-virus (PRNT_80_), and a pseudovirus neutralizing antibody assay (PsVNA_50_).

**Results:** RBD-specific antibodies were already detectable by ACOV2S at the first time point of assessment (d15 after first vaccination), with seroconversion before in all but 2 participants (25 μg dose group); all had seroconverted by Day 29. Across all post-baseline visits, geometric mean concentration of antibody levels were 3.27–7.48-fold higher in the 100 μg compared with the 25 μg dose group. ACOV2S measurements were highly correlated with those from RBD ELISA (Pearson’s r=0.938; p<0.0001) and S-2P ELISA (r=0.918; p<0.0001). For both ELISAs, heterogeneous baseline results and smaller increases in antibody levels following the second vs first vaccination compared with ACOV2S were observed. ACOV2S showed absence of any baseline noise indicating high specificity detecting vaccine-induced antibody response. Moderate–strong correlations were observed between ACOV2S and neutralization tests (nLUC_80_ r=0.933; PsVNA_50_, r=0.771; PRNT_80_, r=0.672; all p≤0.0001).

**Conclusion:** The Elecsys Anti-SARS-CoV-2 S assay (ACOV2S) can be regarded as a highly valuable method to assess and quantify the presence of RBD-directed antibodies against SARS-CoV-2 following vaccination, and may indicate the presence of neutralizing antibodies. As a fully automated and standardized method, ACOV2S could qualify as the method of choice for consistent quantification of vaccine-induced humoral response.

## Introduction

First recognized in Wuhan, China in late 2019, the severe acute respiratory syndrome coronavirus 2 (SARS-CoV-2) has since spread rapidly and infected millions of people globally.(1) The prompt development and approval of vaccines against the virus has been crucial. With over 100 vaccine candidates currently in clinical development,(2) there is a high need for sensitive and specific assays that can reliably quantify immune responses following vaccination.(3)

SARS-CoV-2 is an enveloped positive-sense single-stranded RNA virus containing four structural proteins: spike (S), envelope, membrane, and nucleocapsid (N) protein. The S glycoprotein is proteolytically cleaved into two subunits: S1 containing the host receptor binding domain (RBD) which facilitates entry to host cell through binding to membrane bound angiotensin-converting enzyme 2 (ACE 2), and S2, a membrane-proximal domain.(4) Seroconversion often starts 5–7 days after symptom onset and the antibodies, immunoglobulin M (IgM), IgG and IgA, can be observed after approximately two weeks.(3, 5, 6) While antibody response can be directed against all viral proteins, S and N are considered the main targets of humoral response.(6, 7) Based on the potential for antibodies targeting the spike antigen to inhibit viral entry into the target cells, the majority of vaccine candidates have been designed to induce humoral immune responses against the S antigen.(8) Neutralizing antibodies are important contributors to protective immunity.(3) *In vitro* neutralization testing is a widely applied test to assess the presence of neutralizing antibodies and to titrate them to limiting dilution. A variety of neutralization tests are available, including direct neutralization, which requires biosafety level 3 handling, and pseudotyped-virus assays.(9–11) In convalescent plasma, Ig antibodies towards the SARS-CoV-2 S protein, in particular when directed against the RBD, have been shown to correlate with virus neutralizing titers, suggesting that immunoglobulin levels may predict levels of neutralization.(12, 13) Thus, the potential use of antibody concentrations, quantified by commercially-available immunoassays, as a surrogate for neutralizing titers is currently being explored.(14–16)

The automated, high throughput Roche Elecsys^®^ Anti-SARS-CoV-2 S assay (hereby referred to as ACOV2S) detects and quantifies antibodies against the RBD of the S protein. A previous study showed that the presence of antibodies detected with ACOV2S correlated with the presence of neutralizing antibodies, as detected with direct virus neutralization and surrogate neutralization tests among individuals with minor or no symptoms.(17) In order to generate further supporting evidence for the clinical utility of ACOV2S, we studied the antibody concentration, as measured by ACOV2S, over time in a phase 1 trial of the widely approved, highly effective mRNA-based 2019-nCoV vaccine (mRNA-1273; Moderna, Cambridge, MA) which encodes the stabilized prefusion S trimer, S-2P.(18) We also performed an exploratory analysis comparing ACOV2S results with those from enzyme-linked immunosorbent assays (ELISA) and neutralization tests, based on data from the phase 1 trial.

## Methods

### Study design and participants

We used stored samples from participants enrolled in the phase 1 trial of mRNA-1273 (NCT04283461); full methodological details have previously been described.(18) In this retrospective exploratory analysis, samples from healthy participants aged 18–55 years who received two injections of trial vaccine 28 days apart at a dose of 25 μg or 100 μg were included for assessment. All participants received their first vaccination between March 16 and April 14, 2020.

Blood samples were collected as previously described.(18) Samples collected at baseline (Day 1, first vaccination), and Days 15, 29 (second vaccination), 43 and 57, were analyzed and serum testing was performed at PPD central laboratory (Highland Heights, KY, USA). Informed written consent was originally obtained from all study participants in the context of the associated vaccine phase I study and the study was conducted in accordance with the principles of the Declaration of Helsinki and Good Clinical Practice Guidelines. Approval was granted by the Advarra institutional review board for the phase 1 trial (18) and the diagnostic protocol under which the existing samples were tested.

For comparison of antibody responses induced by vaccination to antibody response to natural SARS-CoV-2 infection, anonymized cross-sectional samples from individuals with polymerase chain reaction (PCR)-confirmed SARS-CoV-2 infection were taken 0–15 days and 16-35 days post-PCR diagnosis and analyzed for presence of SARS-CoV-2 specific antibodies using ACOV2S. These samples were derived from individuals with mild course of disease that underwent quarantine at home or from individuals with more severe course of disease that required hospitalization. All samples were collected between March and July 2020 in Switzerland, Germany, and Ukraine.

### Laboratory assays

#### Elecsys Anti-SARS-CoV-2-S immunoassay (ACOV2S)

The ACOV2S results were measured on a cobas e 602 module (Roche, Mannheim, Germany). All samples were processed according to the manufacturer’s instructions. Measurement results are shown in U/mL, with the cut-off point defined as 0.80 U/mL to differentiate samples as reactive (≥ 0.80 U/mL) and non-reactive (< 0.80 U/mL) for SARS-CoV-2 RBD-specific antibodies. Values between 0.40–250 U/mL represent the linear range. Results below this range were set to 0.4 U/mL and qualified non-reactive. Samples above 250 U/mL were automatically diluted into the linear range of the assay (realized dilutions in this study: 1:10 or 1:100) with Diluent Universal (Roche Diagnostics, Rotkreuz, Switzerland). The analyzer automatically multiplies diluted results with the dilution factor, which in the applied setting enabled an upper limit of quantification of 25000 U/mL for these analyses.

#### Traceability of results to international BAU/mL

Of note, the assigned U/mL are equivalent to Binding Antibody Units (BAU)/mL as defined by the first World Health Organization (WHO) International Standard for anti-SARS-CoV-2 immunoglobulin (NIBSC code 20/136). No conversion of units is required and reported results in U/mL can be directly compared to other studies or results in BAU/mL.

#### Serologic monitoring for breakthrough infections

In addition to the quantification of RBD-specific antibody titers induced by mRNA-1273 vaccination, all samples were also assessed on the same cobas e 602 module with the previously described Elecsys Anti-SARS-CoV-2 immunoassay detecting antibodies to the N protein.(19) As natural infection with SARS-CoV-2 and not vaccination with mRNA-1273 can trigger a positive result in the context of the mRNA-1273 vaccine, this assay was used to determine whether participants were naïve for prior COVID-19 infection or acquired a putative breakthrough infection despite vaccination throughout the period of investigation.

#### Comparator assays

Further assay results were generated under the phase 1 study protocol (18) and the results were transferred to Roche for analysis.

Serum antibody levels against SARS-CoV-2 were measured by ELISA specific to the S protein (stabilized containing 2 Proline mutations and thus referred to as S-2P protein (hereby referred to as S-2P ELISA) and the isolated RBD of the viral S protein (hereby referred to as RBD ELISA). ELISA assay results were expressed as reciprocal endpoint dilution titer. Notably, no reactivity cut-offs or lower limit of quantification were defined and no standardization was applied for either ELISA.

Results from assays that target neutralizing antibodies, providing an estimate of vaccine-induced, antibody-mediated neutralizing activity, were also assessed. These included: 1) a nanoluciferase assay (nLUC) with titers reported as the dilution required to achieve 80% neutralization (80% inhibitory dilution; hereby referred to as nLUC_80_); 2) a live wild-type SARS-CoV-2 plaque reduction neutralization assay (PRNT) with titers expressed as reciprocal of dilution needed for 80% reduction in virus infectivity (hereby referred to as PRNT_80_); and 3) a pseudovirus neutralizing antibody assay (PsVNA) with titers reported as dilution required for 50% neutralization (50% inhibitory dilution; hereby referred to as PsVNA_50_, respectively). Because of the labor-intense nature of the nLUC_80_ and PRNT_80_ assays involving several manual handling steps and cell culture,(18) results were available only for the time points, Day 1, Day 29 (nLUC_80_ only) and Day 43.

Further details of the comparator assay methods have been published.(18, 20) In case no significant inhibition of infection was observed (i.e. < 50% or <80% neutralization) even with the highest sample concentration (i.e. the starting dilution titer), the numerical result of the assay was set to the starting dilution titer and the assay result was interpreted as negative for neutralizing activity in all qualitative concordance analyses. Samples showing significant inhibition (i.e. ≥ 50% or ≥80% neutralization) at any of the applied concentrations were interpreted as positive for neutralizing activity in all qualitative concordance analyses.

### Statistical analysis

For each trial population and dosage group, ACOV2S-measured anti-RBD antibody levels are shown as boxplots (log-scale) for every measurement time point, with values outside the measuring range censored. Comparison of ACOV2S-measured antibody levels per dose group and time point were conducted using reverse cumulative distribution curves. For ACOV2S, geometric mean concentrations (GMCs) and, for ELISA, geometric mean titers (GMTs) were calculated for each time point and stratified by dose group, and 95% confidence intervals (CIs) were calculated by Student’s t distribution on log-transformed data and subsequent back-transformation to original scale.

For the assessment of seroconversion, as measured by ACOV2S, the percentage of subjects who crossed the reactivity cut-off at 0.8 U/mL at or before a given time point was evaluated. A seropositive status was carried forward to later time points.

Pairwise method comparison across all available data points using Passing-Bablok (log-scale) regression analyses (21) with 95% bootstrap CIs were provided for all comparator assays, excluding values outside the measuring range, and Pearson’s correlation coefficients (r) with 95% CIs were calculated.

Qualitative agreement between ACOV2S and neutralization assays was analyzed by positive percentage agreement (PPA), negative percentage agreement (NPA), and overall percentage agreement (OPA), positive and negative predictive value (PPV and NPV) with exact 95% binomial CIs and the positive and negative likelihood ratio with 95% CIs calculated (per Simel et al. approximation (22)). The software R, version 3.4.0, was used for statistical analysis and visualization.(23)

## Results

The analyses included longitudinal sample panels from in total 30 mRNA-1273-vaccinated participants. Of those, 15 participants had received 25 μg dose and the other 15 had received 100 μg dose (both administered as two injections of the indicated dose with a delay of 28 days). Demographics and baseline characteristics of participants have been previously described.(18) In brief, in the 25 μg and 100 μg dose groups, mean ages (±SD) were 36.7 (±7.9) and 31.3 (±8.7) years, 60% and 47% were males, respectively, and the majority were of white ethnicity across both cohorts. All participants were naïve for natural SARS-CoV-2 infection at study start and throughout the investigated timeframe as determined with the Elecsys Anti-SARS-CoV-2 (anti-N) assay (**Supplementary Figure 1**).

### Humoral response after vaccination with mRNA-1273 assessed by Elecsys Anti-SARS-CoV-2 S assay

Anti-RBD antibody levels as measured by ACOV2S, increased over time for both dose groups (**Table 1)**. All participants were non-reactive in ACOV2S at baseline (< 0.4 U/mL), confirming the naïve antibody status for SARS-CoV-2. RBD-specific antibodies were readily detectable by ACOV2S at the first sampling time point (Day 15) and determined high antibody levels indicated that seroconversion had apparently occurred earlier than Day 15 for almost all participants (25 μg: 13/15; 100 μg: 15/15). At Day 29, i.e. day of second vaccination, the remaining two participants of the 25 μg group had seroconverted and developed significant antibody concentrations. The determined antibody concentrations correlated with the applied vaccine dose (**Figure 1A**), with 3.27–7.48-fold higher GMCs observed in the 100 μg group compared with the 25 μg group at all follow up visits (**Table 1**). The 100 μg dose group showed a more homogenous anti-RBD response, as reflected by the smaller geometric standard deviations, indicating reduced inter-individual spread in response to the vaccine at higher dose. In both groups, antibody levels tended to increase until Day 43 and remained high through Day 57 (**Figure 1B**). None of the measured antibody levels exceeded the selected upper limit of quantitation of 25000 U/mL of ACOV2S that resulted from maximally applied 1:100 dilution in this study.

**Figure 1.**
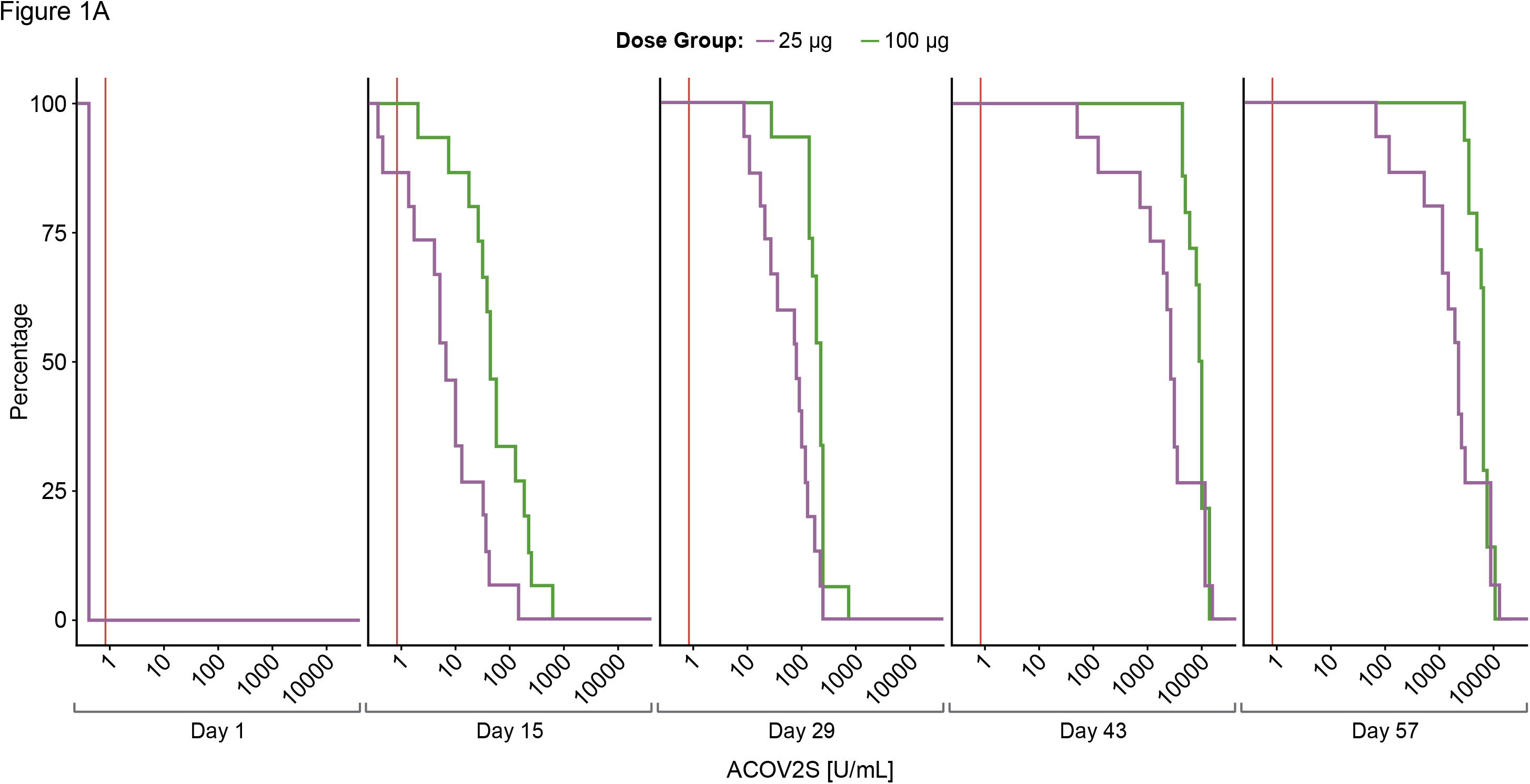
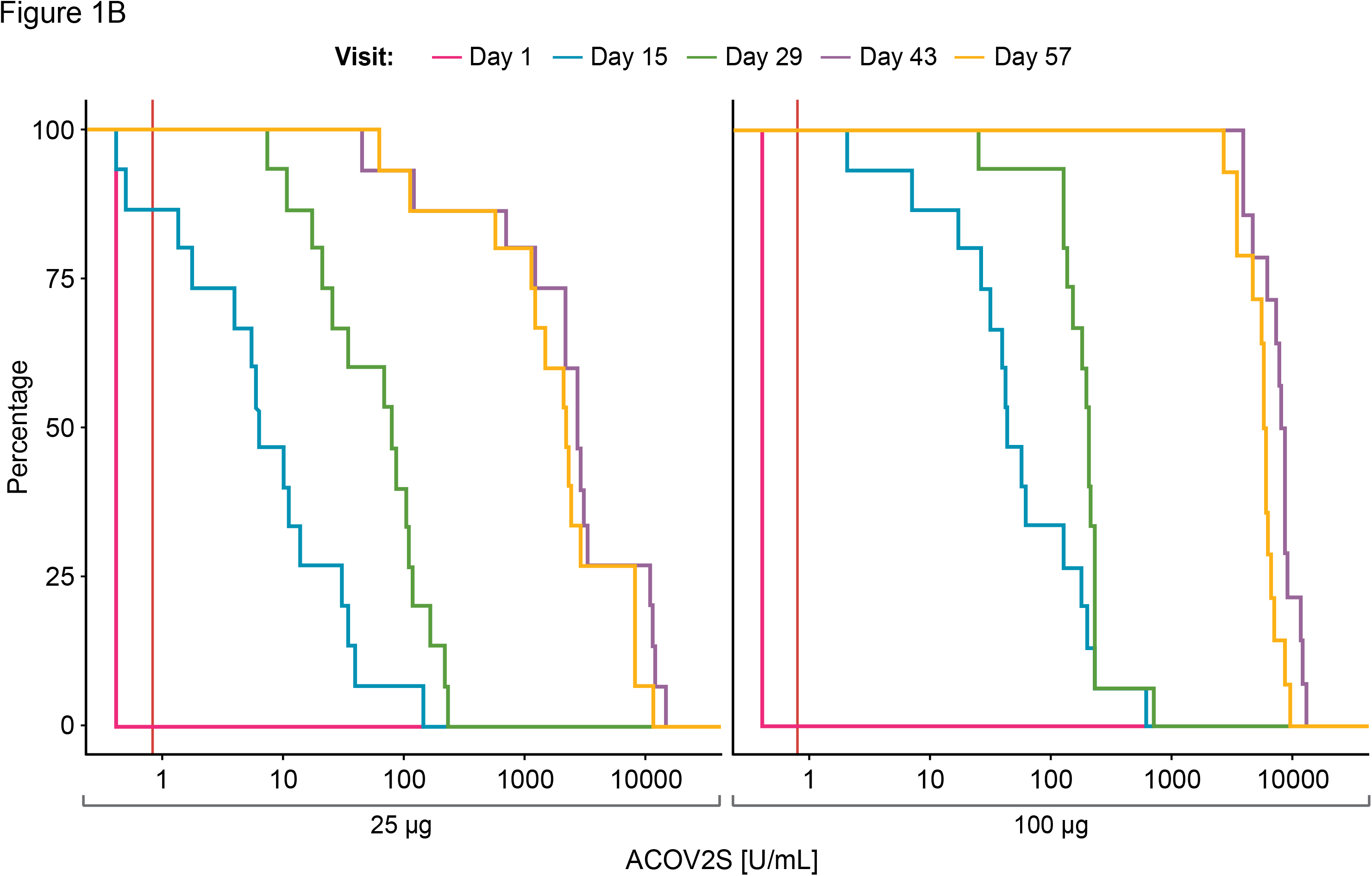
Time-dependent antibody responses as measured by the ACOV2S. Reverse cumulative distribution curves allow for comparison of ACOV2S-measured antibody level distributions between dose groups (Panel A) and visit days (Panel B). Red vertical line indicates reactivity cut-off (0.8 U/mL).

**Table 1.** ACOV2S summary statistics and GMR, comparing the 100 μg group to the 25 μg group.

|  | Day 1 | Day 15 | Day 29 | Day 43 | Day 57 |
| --- | --- | --- | --- | --- | --- |
| **25 µg** | **n=15** | **n=15** | **n=15** | **n=15** | **n=15** |
| **Median** | 0.400 | 6.47 | 79.2 | 2714 | 2176 |
| **Q1–Q3** | 0.400–0.400 | 1.73–30.2 | 21.0–120 | 1156–10918 | 1112–7865 |
| **Min–Max** | 0.400–0.400 | 0.400–147 | 7.49–226 | 45.4–14492 | 62.8–11738 |
| **GMC**<br>**(95% CI)** | 0.400<br>(0.400–0.400) | 6.86<br>(2.75–17.1) | 55.0<br>(30.2–100) | 2123<br>(860–5237) | 1709<br>(745–3920) |
| **GSD** | 1.00 | 5.20 | 2.96 | 5.11 | 4.48 |
| **100 µg** | **n=15** | **n=15** | **n=15** | **n=14** | **n=14** |
| **Median** | 0.400 | 44.7 | 209 | 8476 | 6044 |
| **Q1–Q3** | 0.400–0.400 | 26.9–182 | 135–238 | 6407–9237 | 4637–6998 |
| **Min–Max** | 0.400–0.400 | 2.08–629 | 25.9–726 | 4050–13205 | 2637–9827 |
| **GMC**<br>**(95% CI)** | 0.400<br>(0.400–0.400) | 51.3<br>(23.1–114) | 182<br>(125–264) | 7803<br>(6259–9727) | 5596<br>(4538–6901) |
| **GSD** | 1.00 | 4.21 | 1.97 | 1.46 | 1.44 |
| **GMR (100 vs 25 µg)**<br>**(95% CI)** | 1.00<br>(1.00–1.00) | 7.48<br>(2.35–23.8) | 3.30<br>(1.68–6.49) | 3.68<br>(1.47–9.20) | 3.27<br>(1.41–7.62) |

All values below the lower limit of the measuring range were substituted by this lower limit at 0.4
U/mL. Of note, all ACOV2S levels measured at baseline Day 1 were actually below 0.4 U/mL.
GMC, geometric mean concentrations; GMR, geometric mean ratio; GSD, geometric standard deviation; Q1, first quartile; Q3, third quartile

ACOV2S-measured antibody levels over time in vaccinated samples compared with those in post-PCR confirmed SARS-CoV-2 infection are shown in **Figure 2**. Natively-infected individuals developed a more heterogeneous antibody response to SARS-CoV-2 infection compared with vaccination, likely due to differences in viral load and the course of disease. ACOV2S-measured anti-RBD antibody levels after the first vaccination were within the range developed upon native infection, with levels following the 25 μg dose more aligned with those induced by mild disease (**Figure 2A and 2C**) and the 100 μg dose with severe disease (**Figure 2B and 2D**). After the second vaccination, it can be construed that ACOV2S-measured antibody levels exceeded those induced by native SARS-CoV-2 infection by approximately 10-100 fold.

**Figure 2.**
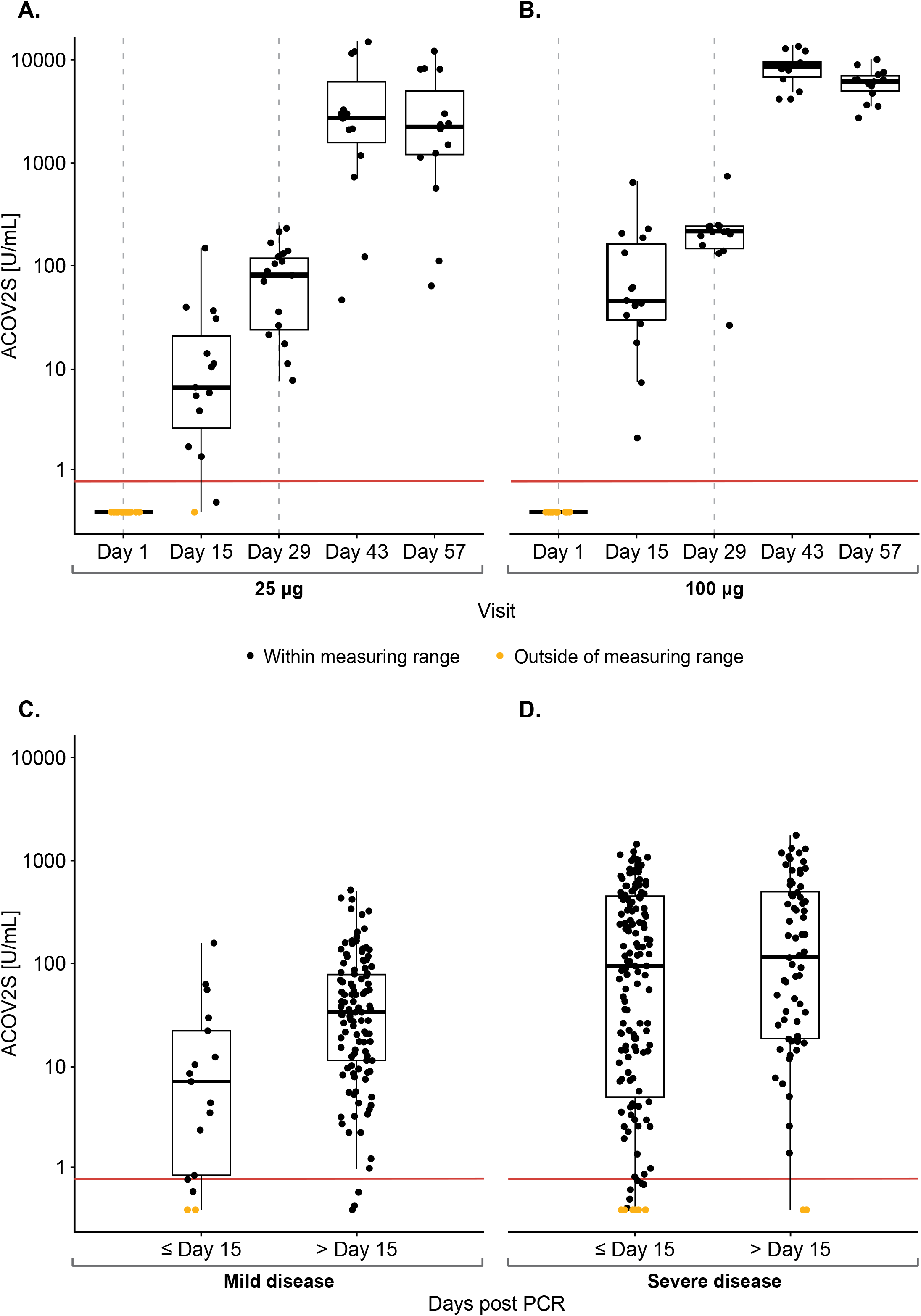
Time course of ACOV2S-measured antibody levels following mRNA-1273 vaccination and native infection. Antibody levels following vaccination are shown in Panel A and B; dotted grey vertical lines indicate time of vaccination, administered at Days 1 and 29. Antibody levels in samples post PCR-confirmed SARS-CoV-2 infection are shown in Panel C and D. Box plots show the individual readouts (black dots) and, 25th, 50th, and 75th percentiles (black box). Red horizontal line indicates reactivity cut-off (0.8 U/mL).

### Concordance of ACOV2S with RBD and S-2P ELISA assays

In total, 113 samples were available for comparative analysis with both ELISA assays across various time points. Measurements by ACOV2S were highly correlated with both RBD ELISA (r=0.938 [95% CI 0.911–0.957]; p<0.0001; **Figure 3A**) and S-2P ELISA (r=0.918 [95% CI 0.883–0.943]; p<0.0001; **Figure 3B**) measurements. Notably, there was distinct heterogeneity of both ELISA results at baseline in contrast to ACOV2S which showed all samples as non-reactive.

**Figure 3.**
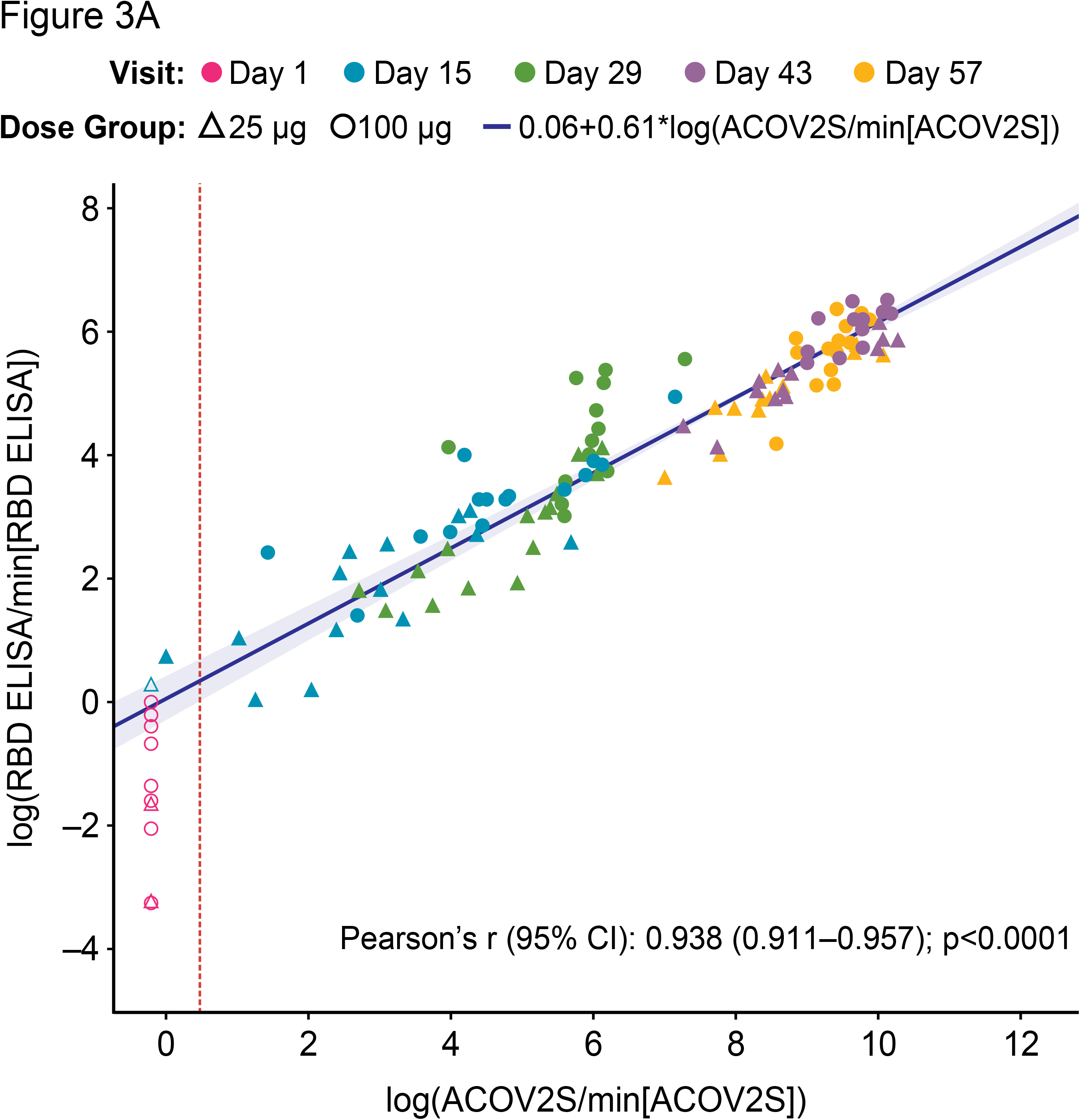
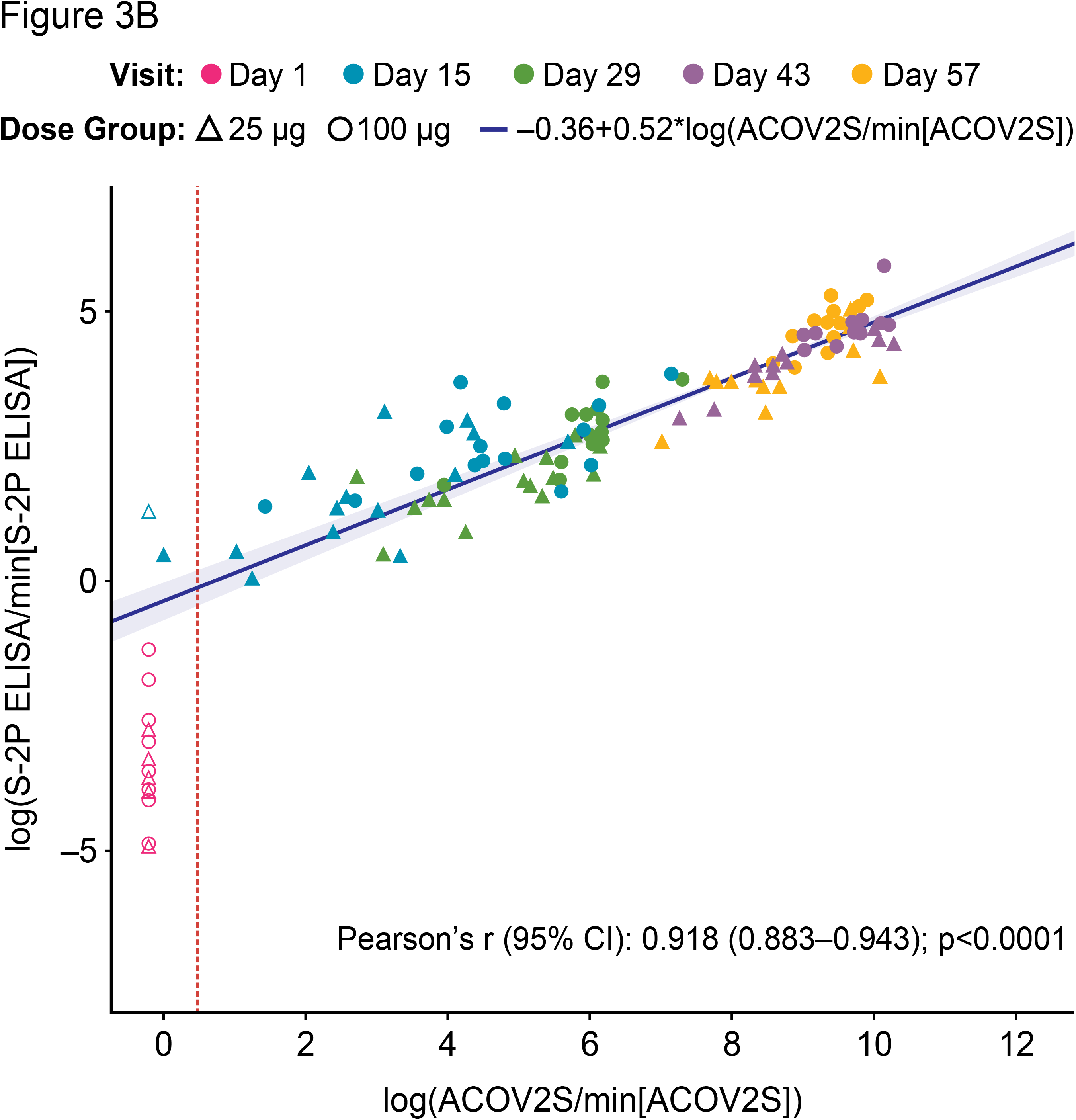
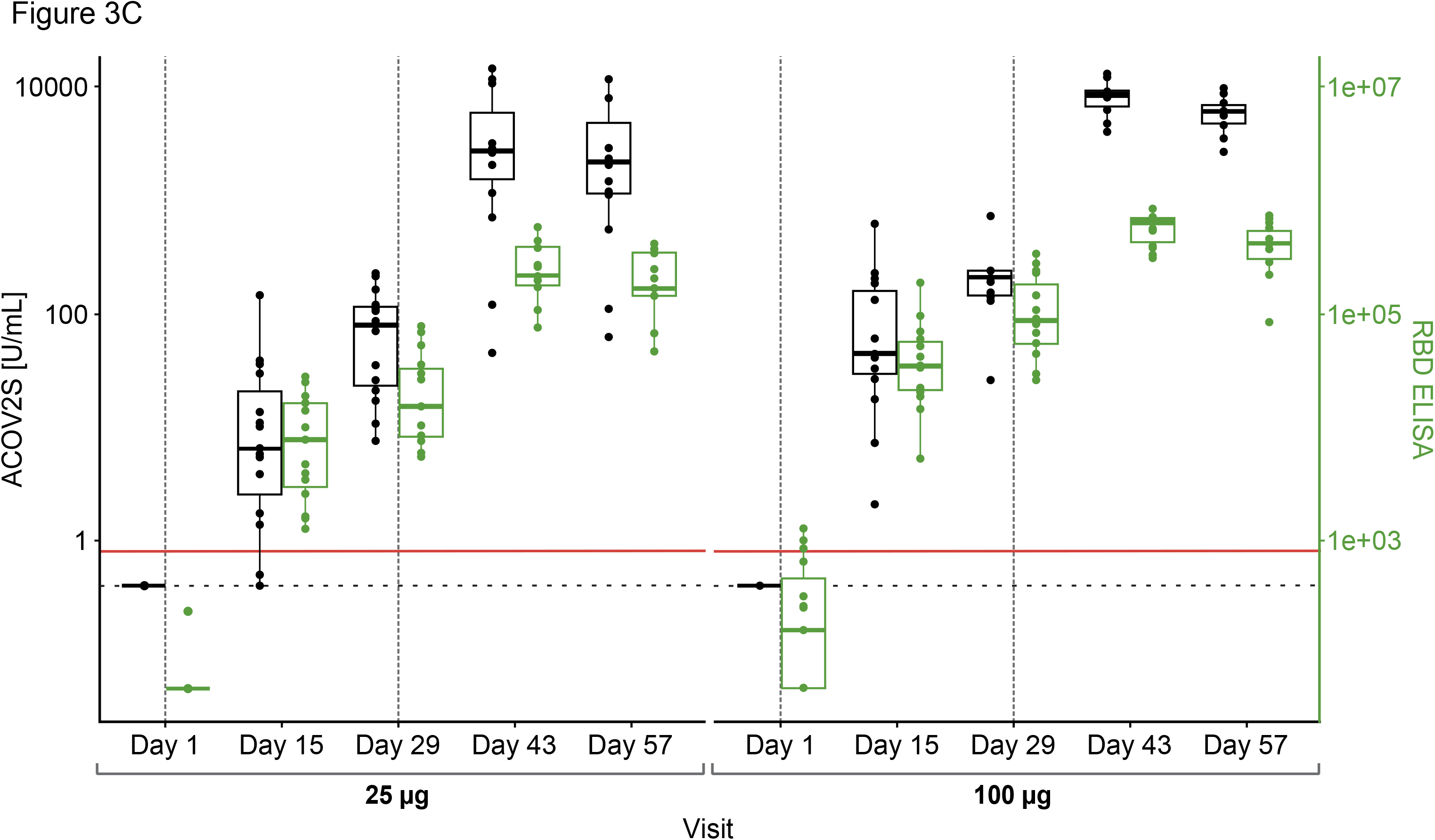
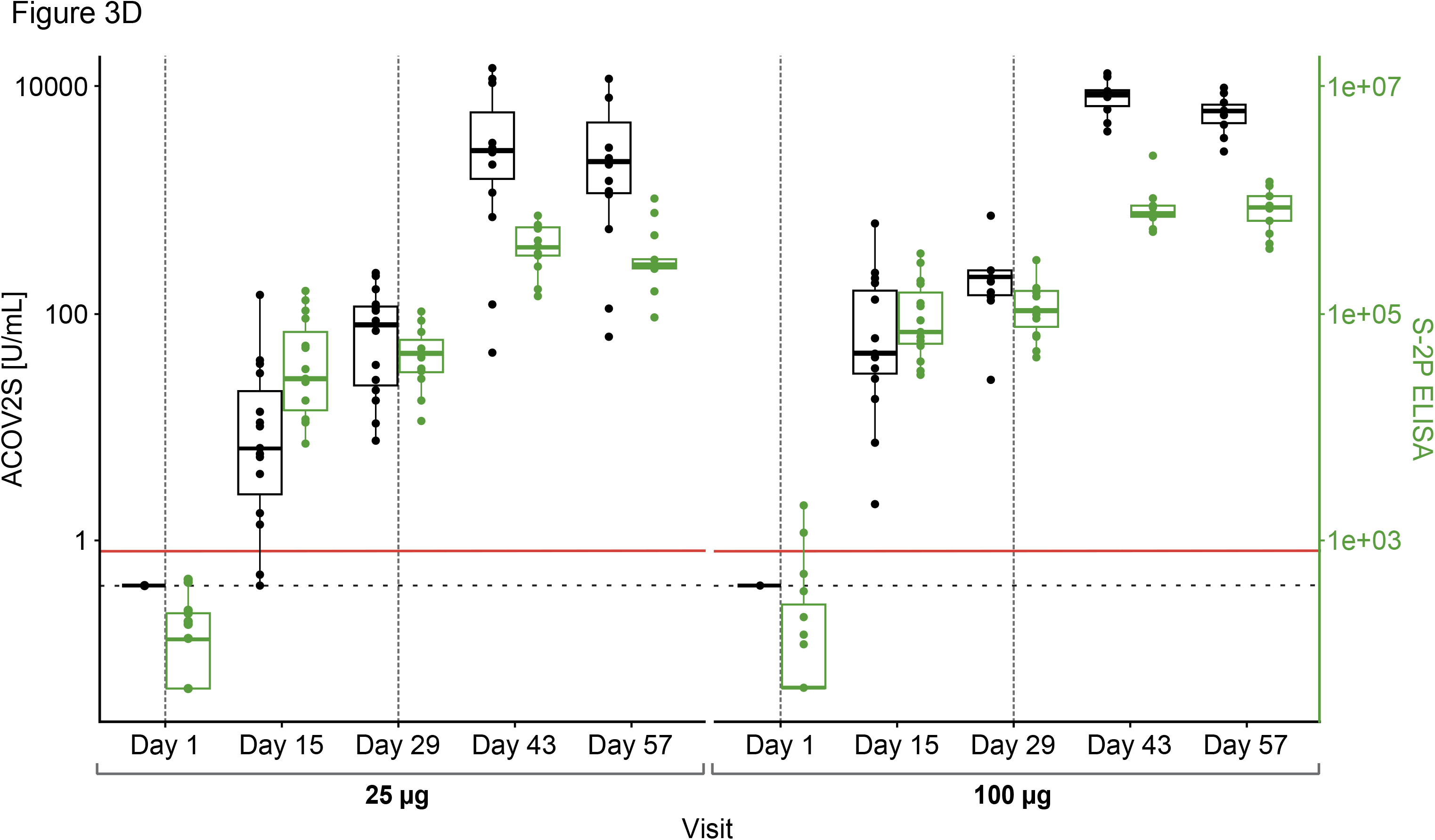
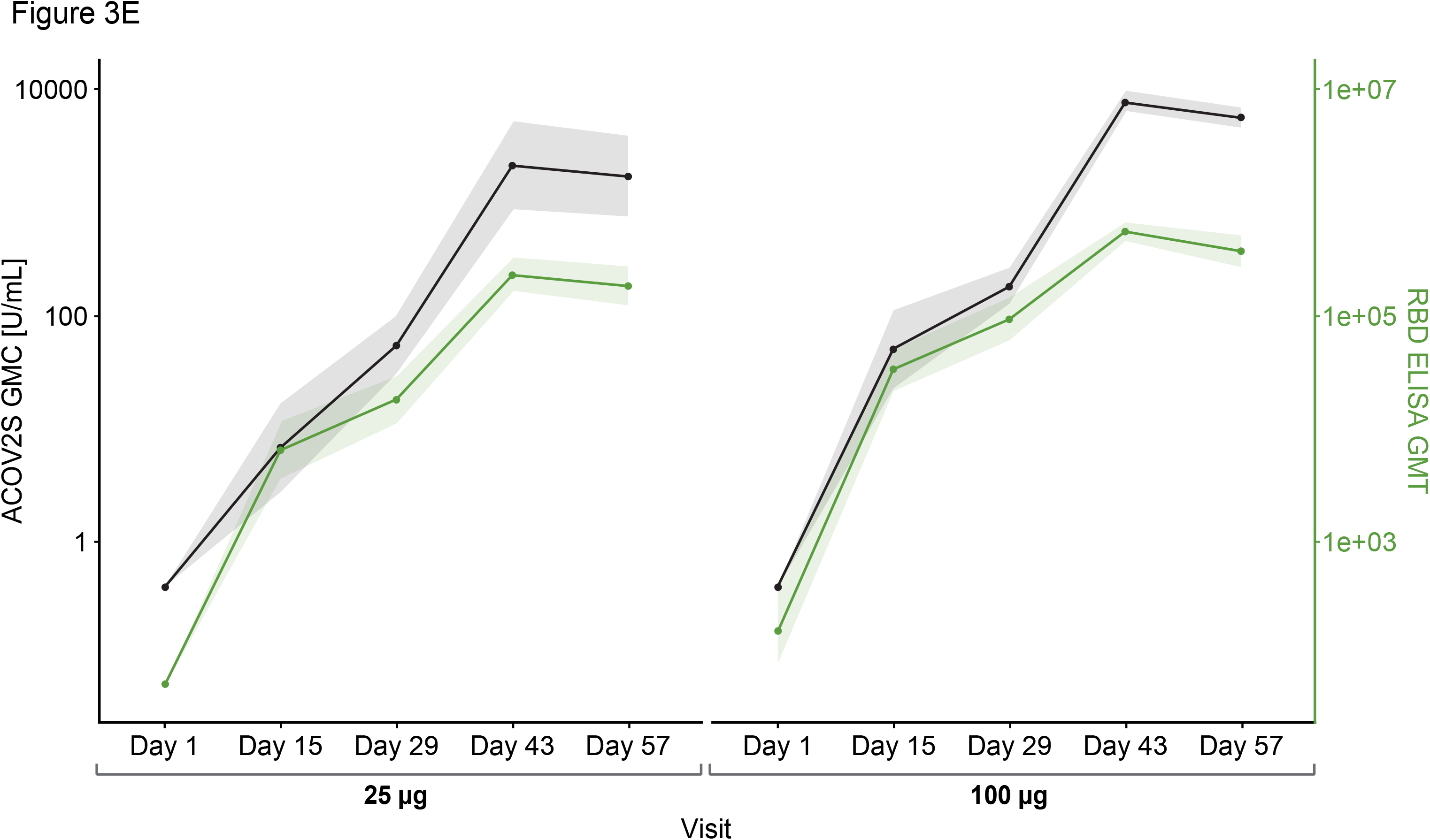
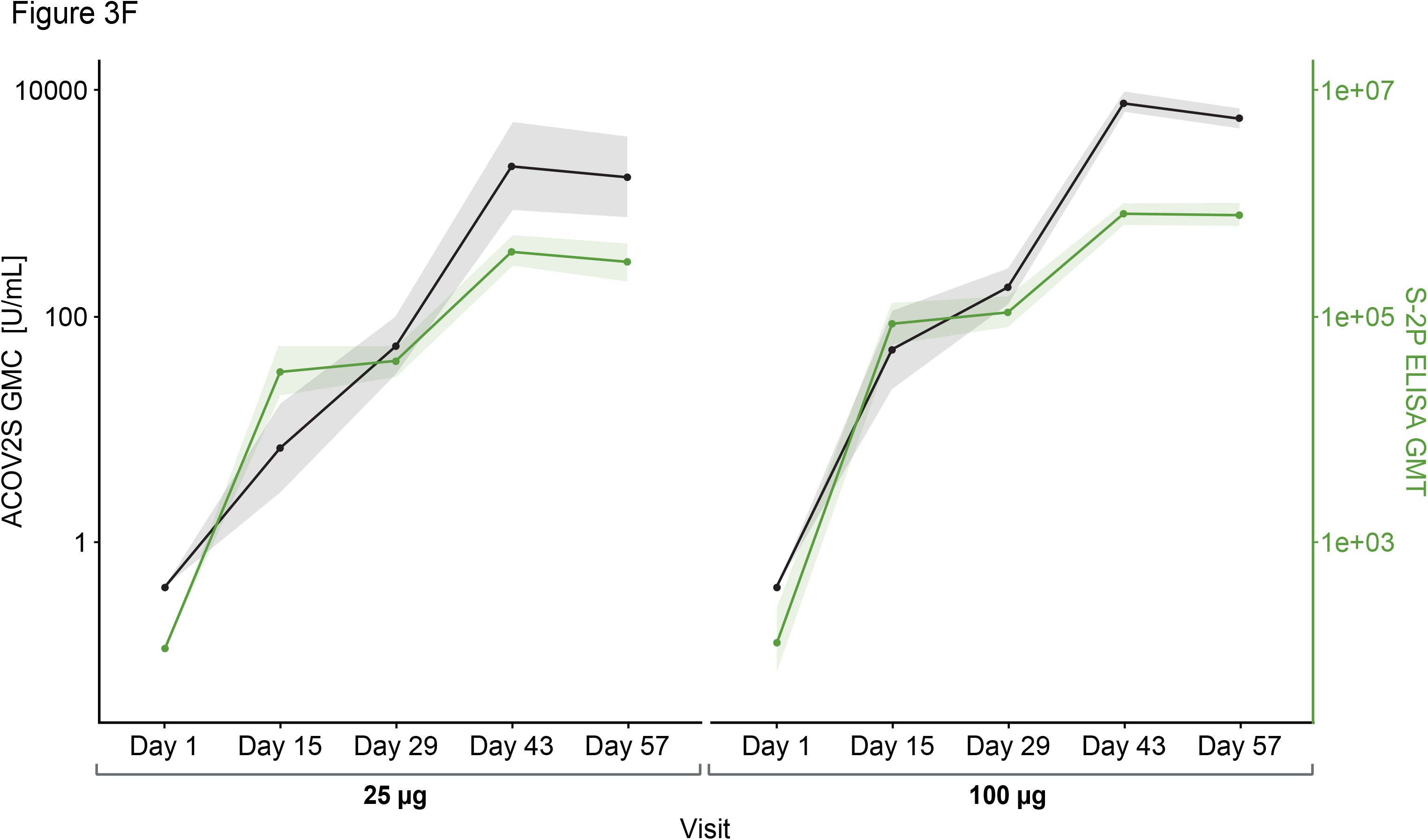
Comparison of ACOV2S and ELISA. Passing–Bablok regression fit (log-scale) for the comparison with RBD ELISA is shown in Panel A, and with S-2P ELISA in Panel B. Red dotted line shows ACOV2S reactivity cut-off. The shaded area represents the 95% confidence interval for the fitted curve. Dots and triangles represent individual samples; filled dots or triangles represent samples within the measuring range for the ACOV2S assay. Time courses of antibody responses measured by RBD ELISA and S-2P ELISA compared to ACOV2S are shown in Panel C and D, respectively. Dotted grey vertical lines show when vaccination injections were administered at Days 1 and 29. Red horizontal line shows ACOV2S reactivity cutoff and the black dashed horizontal line represents the lower end of the ACOV2S measuring range. Box plots show the individual readouts (dots) and, 25th, 50th, and 75th percentiles (box). Time-dependent geometric mean concentrations and geometric mean titers across vaccine dose groups of ACOV2S levels vs RBD ELISA and S-2P ELISA are shown in Panel E and F, respectively. Shaded areas indicate 95% confidence intervals.

Antibody levels measured with ACOV2S and the RBD or S-2P ELISA showed similar time courses following first and second vaccinations (**Figure 3C** and **3D**, respectively). A transient difference became apparent in the 25 μg dose group in which the S-2P ELISA already determined seroconversion for all participants 15 days after first vaccination, whereas ACOV2S did not detect samples from two donors at this time point. A more continuous antibody level development over time and dose groups was observed with the ACOV2S. The obtained results are additionally plotted as GMCs of ACOV2S-measured antibody levels and GMTs of the ELISA-endpoint dilution titers over time in **Figure 3E** and **3F**, respectively, to facilitate relative result comparisons. The ELISA methods showed strong signal increase early after first vaccination followed by a plateau of antibody levels between Day 15 and Day 29, more frequently observed with the S-2P ELISA, and a smaller increase after second vaccination compared with the ACOV2S. This infers that the ELISA methods detect the antibody titer development over time in a more stepwise manner compared to a more continuous antibody titer development as determined with ACOV2S. Also, by making use of the automated onboard dilution, ACOV2S can resolve very high titers while ELISAs appear to approach saturation. This is evident by the more prominent geometric fold-rise after the second vaccination versus the first vaccination for the ACOV2S compared with the ELISA methods (**Supplementary Table 1**).

### Concordance of ACOV2S with neutralization assays

**Figure 4** visualizes concordance of ACOV2S with comparative assays assessing neutralization. For comparison with nLUC_80_, 47 samples had quantifiable results. Numerical correlation with nLUC_80_ measurements was very strong (Pearson’s r=0.933 [95% CI 0.882– 0.962]; p<0.0001) and all samples with a positive nLUC_80_ had a positive ACOV2S measurement (**Figure 4A**). At Day 29, there were 8 samples with a positive ACOV2S result whose nLUC_80_ result was negative, predominantly occurring in 25 μg dose group. A total of 79 samples across all time points had quantifiable PsVNA_50_ results. Strong correlation was observed between ACOV2S and PsVNA_50_ (r=0.771 [0.663–0.848]; p<0.0001) results and all samples with a positive PsVNA_50_ result had a positive ACOV2S measurement. A proportion of samples had a negative PsVNA_50_ result but a positive result with ACOV2S (**Figure 4B**). Analysis with 27 available samples obtained two weeks after the second vaccination (Day 43) showed ACOV2S levels moderately correlated with PRNT_80_ results (r=0.672 [0.392– 0.838]; p=0.0001 [**Figure 4C**]).

**Figure 4.**
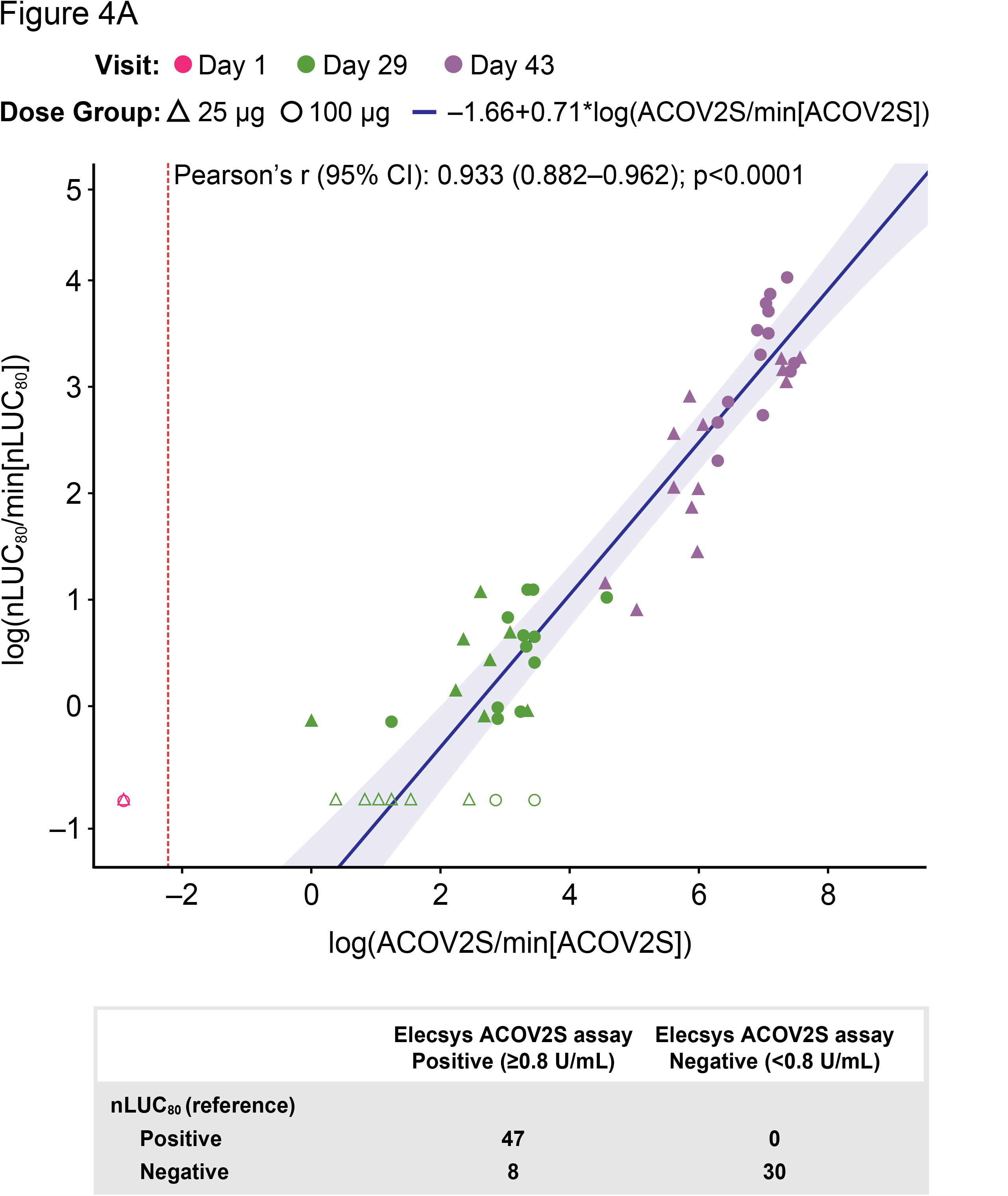
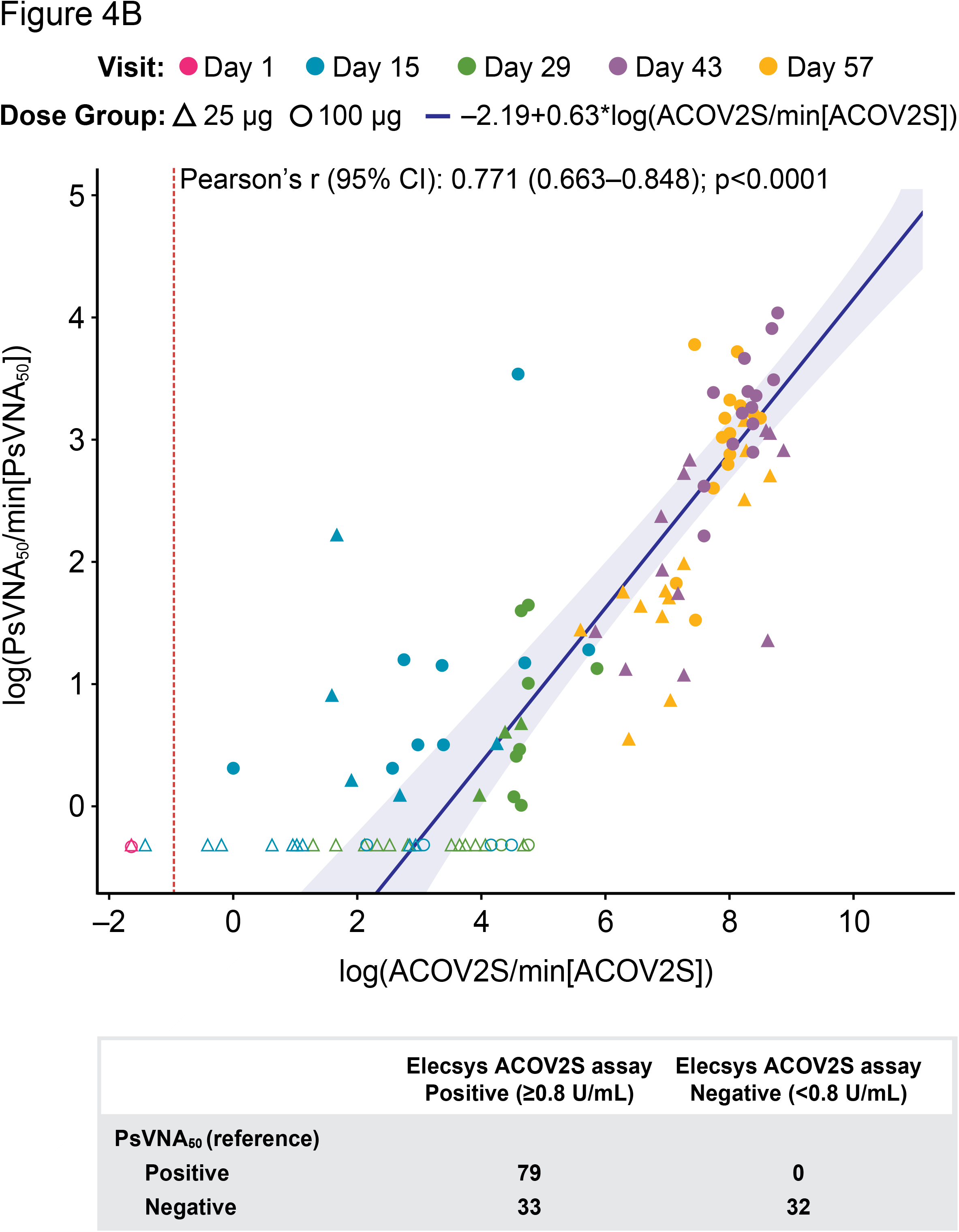
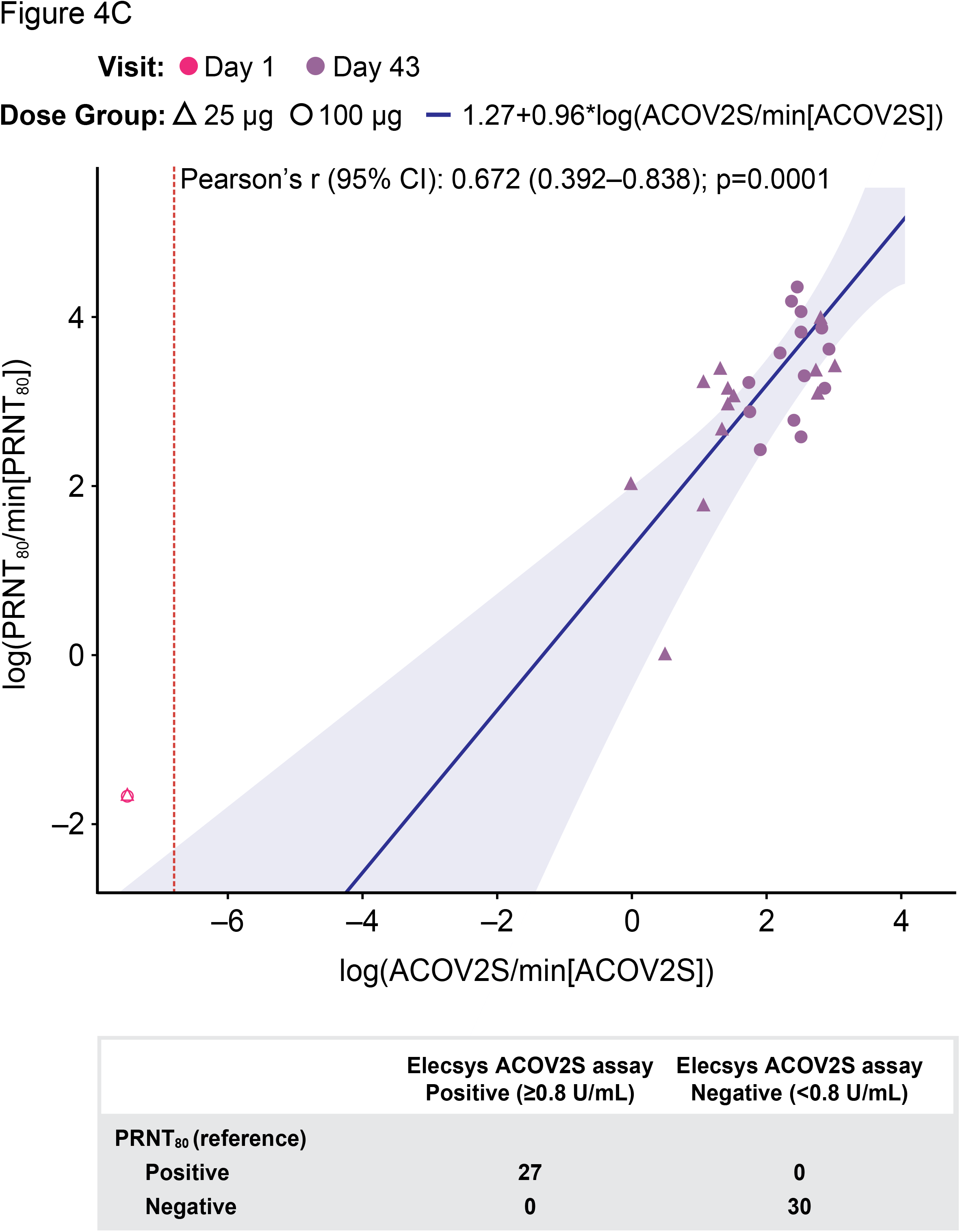
Comparison of ACOV2S and neutralization assays. Passing–Bablok regression fit (log-scale) of ACOV2S with nLUC_80_ is shown in Panel A, PsVNA_50_ in Panel B and PRNT_80_ in Panel C. Red dotted line shows ACOV2S reactivity cut-off. The shaded area represents the 95% confidence interval for the fitted curve. Dots or crosses show individual sample readouts. Filled dots or triangles represent samples within the measuring range for both ACOV2S and respective comparator assay. Overlaid table shows the qualitative agreement between Elecsys ACOV2S and comparator assays.

Qualitative agreement between ACOV2S and neutralizing test results is presented in **Supplementary Table 2**. The PPA and NPV for all neutralization assays was 100%, highlighting that no samples were negative for ACOV2S while positive for neutralization.

## Discussion

Immune responses to SARS-CoV-2 infection and vaccination can significantly vary with each individual(24–26) and longevity of the humoral immune response to SARS-CoV-2 has repeatedly been a matter of investigation.(27) Correlation of protection from symptomatic disease with determined antibody titers is also being explored.(28) Here, reliable correlation requires evaluation of large cohorts and multi-centric datasets and determination of titers with a standardized and globally available method. The reported high efficacy of the mRNA1273 vaccine renders breakthrough infections rare and non-responders unlikely.(18, 29, 30) Together with the rapidly growing number of SARS-CoV-2 vaccines in development, these aspects further emphasize the need for automated, high-throughput methods to reliably quantify immune response in a standardized manner to enable large dataset comparisons, confirm seroconversion in all targeted individuals, independent of pre-existing conditions or medications,(31) as well as long-term monitoring.

In this exploratory analysis of mRNA-1273-vaccinated human samples from the phase 1 trial,(18) the quantification of the anti-RBD antibodies through ACOV2S allowed the monitoring of changes between visits and resolution of differences between dosage groups, with antibody concentrations increasing in a time- and dose-dependent manner. Primary vaccination resulted in seroconversion in all participants early after the first injection. Seroconversion after initial vaccination and overall anti-RBD concentration development after application of 100 μg per injection was stronger than 25 μg. Antibody levels present two weeks after second vaccination with mRNA-1273 were seen to exceed those induced by natural SARS-CoV-2 infection, both of which provide protection against symptomatic infection with higher antibody levels expected to be synonymous with longevity.(29, 32, 33) Results from ACOV2S compared well to those obtained with two ELISA methods, one targeting antibodies against the S-2P protein of the virus and the other specifically against the RBD domain (both r>0.9; p<0.0001). However, there was high heterogeneity in ELISA baseline values, potentially due to less specific signals. The lower end of measuring range is not defined for either ELISA. Additionally, no validated reactivity cut-off was available, hence it was not possible to formally assess the qualitative agreement between the ACOV2S and ELISA methods. A more continuous increase of titer up to peak was observed with the ACOV2S, while the ELISA measurements seemed to approach a saturation limit. Of note, the linear range and thus the upper limit of quantitation has not been established for either ELISA. Despite our findings and previous studies suggesting that S-focused ELISAs may offer greater sensitivity,(34) antibody responses measured with the RBD ELISA were similar to the S-2P ELISA, with better signal dynamics illustrated by the more homogenous increase in GMT. Additionally, the high S-2P ELISA titers detected soon after the first vaccination, even with the low 25 μg dose, could misleadingly be interpreted as suggestive of strong immune response from early on, while efficient immunity has been reported to occur only later after vaccination.(30) In contrast, dynamic increase of antibody levels accompanying vaccination enable better characterization of the developing immune response than plateau reactivity. Less variation in baseline titer was also observed with the RBD ELISA, potentially due to lower cross-reactivity with antibodies to previously endemic and highly abundant coronavirus strains, which show structural similarities in the S2 subunit.(35) Taking into consideration that the RBD is poorly conserved among them,(7) antibody-detection assays specifically targeting antibodies directed against the RBD appear highly suitable for quantifying the humoral immune response to SARS-CoV-2.

In this study, good correlation was observed with ACOV2S and the established surrogate neutralization tests, nLUC_80,_ and PsVNA_50_. Disagreement was observed only with earlier samples where some positive, but relatively low ACOV2S results coincided with non-reactive neutralizing antibody test results. This was possibly due to insufficient antibody concentrations to prevent infection in the *in vitro* setting of a neutralization test, supporting the clinical finding that single dose vaccination does not convey optimal protection from infection and that two-step vaccination inducing higher antibody titers is required. With an apparent more continuous resolution of antibody development, these observations suggest ACOV2S might allow for more precise timing of reaching putatively protective levels than methods with rapidly developing plateaus. Although limited to samples from a singular visit (Day 43), we found ACOV2S levels also correlated with live-virus neutralization test PRNT_80_ titers. For all three neutralization tests, appearance of neutralizing effects was suggested within two weeks of the second vaccination, further supporting the need for a two-dose schedule. Also, it has been described manifold that RBD is not necessarily the exclusive, yet the dominant target for antibody-mediated virus neutralization, meaning that RBD-directed antibodies contribute to virus neutralization. Together with the observed rapid development of very high anti-RBD titers illustrating the strong immunogenic potential of the mRNA-1273 vaccine especially with the clinically-selected 100 μg dose,(29) these findings render anti-RBD levels a suitable and convenient surrogate marker for the presence of neutralizing antibodies during vaccination monitoring, with high levels suggestive of greater protective immunity.

Live virus neutralization using wild type virus requires handling of live SARS-CoV-2 in a specialized biosafety level 3 containment facility and is time-consuming, deeming it unsuitable for large scale use. Neutralization test methods using replication-defective pseudotyped viral particles have been developed; however, these still require live-cell culture, considerable manual handling steps and, consequently, inevitable variance in neutralization results. Although surrogate neutralization assays have been developed and validated,(10, 36) their applied competitive assay principle goes along with a rather small dynamic range, which limits resolution of change in high titer vaccination samples. In addition, challenges of semi-automatable methods and costs remain. Also, neutralization tests are potentially limited in that they only address static antibody levels at a given time point and do not take into account antibody avidity, maturation or the immediate restimulation of the immune memory by a recurring infection *in vivo*. Poor signal resolution at the lower end of the measuring range of neutralization tests is also a concern. The ACOV2S assay has been developed to detect the presence of low levels of RBD-directed antibodies with a high sensitivity (97.92%; 95% CI: 95.21–99.32) and specificity (99.95%; 95% CI: 99.87–99.99),(37) and a medical decision point at 0.8 U/mL as an indicator of infection or vaccination, i.e. the lowest quantity of antibody that determines reactivity for SARS-CoV-2 RBD-specific antibodies. The quantitative setup of the assay however allows for definition of additional medical decision points that might best suit other purposes, like protective levels of antibody or high titer plasma for therapeutic use.(38) In addition, ACOV2S is standardized congruently to the first WHO international standard and assigned units can be used interchangeably to BAU/mL, making it suitable for long-term monitoring and referencing of results to the international standard.

Altogether, these findings suggest that ACOV2S levels may predict the presence of neutralizing antibodies,(17) especially at later time points after vaccination, and therefore, potentially provide a more accessible method for enumerating immune response in vaccinated individuals. However, ongoing research is required to elucidate if protective anti-RBD thresholds can be defined that are indicative of, for example, sterile immunity or of preventing symptomatic infection.

Limitations of our study include the small sample size as well as the lack of variation in time points available for analysis for some of the neutralization assays. The relatively short follow-up mitigates analysis of the ability of ACOV2S assay to determine the sustainability of antibody response. Further comparison studies using longer term follow-up and bigger samples sizes are warranted.

## Conclusion

Assessing the longevity of antibody titers over time together with monitoring for symptomatic re-infection is essential to determine long-term immune protection and define antibody levels as a reliable and conveniently accessible surrogate marker of protection. These data indicate that the ACOV2S immunoassay can be regarded as a highly valuable, convenient and widely accessible method to assess and quantify the presence of antibodies directed at the RBD of SARS-CoV-2, conducive to immune response. ACOV2S sensitively detects and reliably quantifies the vaccination-induced humoral response over a dynamic range that can be conveniently scaled by automated onboard dilution. Our results support the potential for RBD-based immunoassays to replace neutralization tests in the assessment of immune response after vaccination against SARS-CoV-2. These findings also support the use of ACOV2S for longitudinal response monitoring of the RBD-specific antibody response to vaccination and, ultimately, the investigation of an antibody-based correlate of protection from symptomatic COVID-19.

## Data Availability

The authors are committed to sharing data supporting the findings of eligible studies. Access to de-identified patient-level data and supporting clinical documents with qualified external researchers may be available upon request once the trial is complete.

## Declarations

### Funding

This study utilized samples obtained under NCT04283461 funded by NIAID, National Institutes of Health (NIH), Bethesda MD USA. Moderna provided mRNA-1273 samples from their phase 1 trial for this study but did not provide any financial support. The phase 1 trial was supported by the NIAID, National Institutes of Health (NIH), Bethesda MD USA, under award numbers UM1AI148373 (Kaiser Washington), UM1AI148576 (Emory University), UM1AI148684 (Emory University), UM1Al148684-01S1 (Vanderbilt University Medical Center), and HHSN272201500002C (Emmes); by the National Center for Advancing Translational Sciences, NIH, under award number UL1 TR002243 (Vanderbilt University Medical Center); and by the Dolly Parton COVID-19 Research Fund (Vanderbilt University Medical Center).This analysis was funded by Roche Diagnostics GmbH (Penzberg, Germany). Editorial support was provided by Jade Drummond of inScience Communications, Springer Healthcare Ltd, UK, and was funded by Roche Diagnostics International Ltd (Rotkreuz, Switzerland).

### Competing interests

Simon Jochum, Imke Kirste, Sayuri Hortsch, Veit Peter Grunert and Udo Eichenlaub are employees of Roche Diagnostics. Basel Kashlan is an employee of PPD, Inc. Holly Legault and Rolando Pajon are employees of Moderna, Inc.

## Acknowledgements

We would like to thank the associated mRNA-1273 Study Team for their contribution to data collection as part of the associated Phase I study (NCT04283461). Additionally, we would like to thank Micah Taylor, Kristin Lucas and the lab operators at PPD Central Labs (Kentucky, USA) for their contribution to data collection and study execution. We would also like to acknowledge and thank Celine Leroy, Tara Pigg, Emma Tao, Yuli Sun, Walter Stoettner (Roche Diagnostics) and Maha Maglinao (Moderna) for their individual contributions to study execution. Additional gratitude goes to Laura Schlieker (Staburo GmbH) for the validation of the analysis. ELECSYS and COBAS are trademarks of Roche. All other product names and trademarks are the property of their respective owners. Assessment of humoral response following vaccination is currently outside of the intended use of the Elecsys Anti-SARS-CoV-2 S assay.

## Author contributions

Study concept/design: IK, UE, SJ, SH

Data acquisition: IK, BK

Data analysis and interpretation: SH, VPG, SJ, IK

Review and final approval of the manuscript: SJ, IK, SH, HL, RP

## Notes

### Clinical Trial

NCT04283461

### Author Declarations

Approval was granted by the Advarra institutional review board for the phase 1 trial and the diagnostic protocol under which the existing samples were tested.

### Summary of Updates

Figures updated to include labels

## References

1. Center JHUCR. COVID-19 dashboard by the Center for Systems Science and Engineering (CSSE) at Johns Hopkins University. 2021 [Available from: https://coronavirus.jhu.edu/map.html.

2. COVID-19 W. COVID-19 vaccine tracker and landscape 2021 [Available from: https://www.who.int/publications/m/item/draft-landscape-of-covid-19-candidate-vaccines.

3. Carrillo J, Izquierdo-Useros N, Avila-Nieto C, Pradenas E, Clotet B, Blanco J. Humoral immune responses and neutralizing antibodies against SARS-CoV-2; implications in pathogenesis and protective immunity. Biochem Biophys Res Commun. 2021;538:187–91.

4. Walls AC, Park YJ, Tortorici MA, Wall A, McGuire AT, Veesler D. Structure, Function, and Antigenicity of the SARS-CoV-2 Spike Glycoprotein. Cell. 2020;183(6):1735.

5. Lou B, Li TD, Zheng SF, Su YY, Li ZY, Liu W, et al. Serology characteristics of SARS-CoV-2 infection after exposure and post-symptom onset. Eur Respir J. 2020;56(2).

6. Zhao J, Yuan Q, Wang H, Liu W, Liao X, Su Y, et al. Antibody Responses to SARS-CoV-2 in Patients With Novel Coronavirus Disease 2019. Clin Infect Dis. 2020;71(16):2027–34.

7. Premkumar L, Segovia-Chumbez B, Jadi R, Martinez DR, Raut R, Markmann A, et al. The receptor binding domain of the viral spike protein is an immunodominant and highly specific target of antibodies in SARS-CoV-2 patients. Sci Immunol. 2020;5(48).

8. Dai L, Gao GF. Viral targets for vaccines against COVID-19. Nat Rev Immunol. 2021;21(2):73–82.

9. Bewley KR, Coombes NS, Gagnon L, McInroy L, Baker N, Shaik I, et al. Quantification of SARS-CoV-2 neutralizing antibody by wild-type plaque reduction neutralization, microneutralization and pseudotyped virus neutralization assays. Nat Protoc. 2021;16(6):3114–40.

10. Tan CW, Chia WN, Qin X, Liu P, Chen MI, Tiu C, et al. A SARS-CoV-2 surrogate virus neutralization test based on antibody-mediated blockage of ACE2-spike protein-protein interaction. Nat Biotechnol. 2020;38(9):1073–8.

11. Riepler L, Rossler A, Falch A, Volland A, Borena W, von Laer D, et al. Comparison of Four SARS-CoV-2 Neutralization Assays. Vaccines (Basel). 2020;9(1).

12. Legros V, Denolly S, Vogrig M, Boson B, Siret E, Rigaill J, et al. A longitudinal study of SARS-CoV-2-infected patients reveals a high correlation between neutralizing antibodies and COVID-19 severity. Cell Mol Immunol. 2021;18(2):318–27.

13. Salazar E, Kuchipudi SV, Christensen PA, Eagar T, Yi X, Zhao P, et al. Convalescent plasma anti-SARS-CoV-2 spike protein ectodomain and receptor-binding domain IgG correlate with virus neutralization. J Clin Invest. 2020;130(12):6728–38.

14. Bal A, Pozzetto B, Trabaud MA, Escuret V, Rabilloud M, Langlois-Jacques C, et al. Evaluation of High-Throughput SARS-CoV-2 Serological Assays in a Longitudinal Cohort of Patients with Mild COVID-19: Clinical Sensitivity, Specificity, and Association with Virus Neutralization Test. Clin Chem. 2021;67(5):742–52.

15. Irsara C, Egger AE, Prokop W, Nairz M, Loacker L, Sahanic S, et al. Clinical validation of the Siemens quantitative SARS-CoV-2 spike IgG assay (sCOVG) reveals improved sensitivity and a good correlation with virus neutralization titers. Clin Chem Lab Med. 2021;59(8):1453–62.

16. Padoan A, Bonfante F, Pagliari M, Bortolami A, Negrini D, Zuin S, et al. Analytical and clinical performances of five immunoassays for the detection of SARS-CoV-2 antibodies in comparison with neutralization activity. EBioMedicine. 2020;62:103101.

17. Rubio-Acero R, Castelletti N, Fingerle V, Olbrich L, Bakuli A, Wolfel R, et al. In Search of the SARS-CoV-2 Protection Correlate: Head-to-Head Comparison of Two Quantitative S1 Assays in Pre-characterized Oligo-/Asymptomatic Patients. Infect Dis Ther. 2021:1–14.

18. Jackson LA, Anderson EJ, Rouphael NG, Roberts PC, Makhene M, Coler RN, et al. An mRNA Vaccine against SARS-CoV-2 - Preliminary Report. N Engl J Med. 2020;383(20):1920–31.

19. Muench P, Jochum S, Wenderoth V, Ofenloch-Haehnle B, Hombach M, Strobl M, et al. Development and Validation of the Elecsys Anti-SARS-CoV-2 Immunoassay as a Highly Specific Tool for Determining Past Exposure to SARS-CoV-2. J Clin Microbiol. 2020;58(10).

20. Anderson EJ, Rouphael NG, Widge AT, Jackson LA, Roberts PC, Makhene M, et al. Safety and Immunogenicity of SARS-CoV-2 mRNA-1273 Vaccine in Older Adults. N Engl J Med. 2020;383(25):2427–38.

21. Passing H, Bablok. A new biometrical procedure for testing the equality of measurements from two different analytical methods. Application of linear regression procedures for method comparison studies in clinical chemistry, Part I. J Clin Chem Clin Biochem. 1983;21(11):709–20.

22. Simel DL, Samsa GP, Matchar DB. Likelihood ratios with confidence: sample size estimation for diagnostic test studies. J Clin Epidemiol. 1991;44(8):763–70.

23. Team RC. R: A language and environment for statistical computing. R Foundation for Statistical Computing, Vienna, Austria. 2020 [Available from: https://www.R-project.org/.

24. Brodin P, Jojic V, Gao T, Bhattacharya S, Angel CJ, Furman D, et al. Variation in the human immune system is largely driven by non-heritable influences. Cell. 2015;160(1-2):37–47.

25. Castro Dopico X, Ols S, Lore K, Karlsson Hedestam GB. Immunity to SARS-CoV-2 induced by infection or vaccination. J Intern Med. 2021.

26. Jonsson S, Sveinbjornsson G, de Lapuente Portilla AL, Swaminathan B, Plomp R, Dekkers G, et al. Identification of sequence variants influencing immunoglobulin levels. Nat Genet. 2017;49(8):1182–91.

27. Turner JS, Kim W, Kalaidina E, Goss CW, Rauseo AM, Schmitz AJ, et al. SARS-CoV-2 infection induces long-lived bone marrow plasma cells in humans. Nature. 2021;595(7867):421–5.

28. Gaebler C, Wang Z, Lorenzi JCC, Muecksch F, Finkin S, Tokuyama M, et al. Evolution of antibody immunity to SARS-CoV-2. Nature. 2021;591(7851):639–44.

29. Baden LR, El Sahly HM, Essink B, Kotloff K, Frey S, Novak R, et al. Efficacy and Safety of the mRNA-1273 SARS-CoV-2 Vaccine. N Engl J Med. 2021;384(5):403–16.

30. Chu L, McPhee R, Huang W, Bennett H, Pajon R, Nestorova B, et al. A preliminary report of a randomized controlled phase 2 trial of the safety and immunogenicity of mRNA-1273 SARS-CoV-2 vaccine. Vaccine. 2021;39(20):2791–9.

31. Rubbert-Roth A, Vuilleumier N, Ludewig B, Schmiedeberg K, Haller C, von Kempis J. Anti-SARS-CoV-2 mRNA vaccine in patients with rheumatoid arthritis. Lancet Rheumatol. 2021;3(7):e470–e2.

32. Feng C, Shi J, Fan Q, Wang Y, Huang H, Chen F, et al. Protective humoral and cellular immune responses to SARS-CoV-2 persist up to 1 year after recovery. Nat Commun. 2021;12(1):4984.

33. Figueiredo-Campos P, Blankenhaus B, Mota C, Gomes A, Serrano M, Ariotti S, et al. Seroprevalence of anti-SARS-CoV-2 antibodies in COVID-19 patients and healthy volunteers up to 6 months post disease onset. Eur J Immunol. 2020;50(12):2025–40.

34. Tian Y, Lian C, Chen Y, Wei D, Zhang X, Ling Y, et al. Sensitivity and specificity of SARS-CoV-2 S1 subunit in COVID-19 serology assays. Cell Discov. 2020;6:75.

35. Ladner JT, Henson SN, Boyle AS, Engelbrektson AL, Fink ZW, Rahee F, et al. Epitope-resolved profiling of the SARS-CoV-2 antibody response identifies cross-reactivity with endemic human coronaviruses. Cell Rep Med. 2021;2(1):100189.

36. Marien J, Michiels J, Heyndrickx L, Nkuba-Ndaye A, Ceulemans A, Bartholomeeusen K, et al. Evaluation of a surrogate virus neutralization test for high-throughput serosurveillance of SARS-CoV-2. J Virol Methods. 2021:114228.

37. Riester E, Findeisen J, Hegel K, Kabesch M, Ambrosch A, Rank CM, et al. Performance evaluation of the Roche Elecsys Anti-SARS-CoV-2 S immunoassay medRxiv 2021 [Available from: https://www.medrxiv.org/content/10.1101/2021.03.02.21252203v1.

38. Hinton DM. Convelescent Plasma EUA Letter of Authorization: FDA; 9 March 2021 [Available from: https://www.fda.gov/media/141477/download.

