## Supplementary material for "Clinical utility of Elecsys Anti-SARS-CoV-2 S assay in COVID-19 vaccination: An exploratory analysis of the mRNA-1273 phase 1 trial"

**Supplementary Table 1. Geometric mean-fold rises over time with ACOV2S and ELISA methods.**

|  | **Day 1** | **Day 15** | **Day 29** | **Day 43** | **Day 57** |
| --- | --- | --- | --- | --- | --- |
| **25 μg** | | | | | |
| **RBD ELISA**  **N**  **GMFR**  **(95% CI)** | 15  1.00  (1.00–1.00) | 15  118  (60.9–230) | 15  327  (195–549) | 13  4133  (2715–6291) | 13  3254  (1990–5231) |
| **S-2P ELISA**  **N**  **GMFR**  **(95% CI)** | 15  1.00  (1.00–1.00) | 15  279  (121–644) | 15  348  171–708) | 13  2855  (1490–5588) | 13  2278  (1062–4885) |
| **ACOV2S**  **N**  **GMFR**  **(95% CI)** | 15  1.00  (1.00–1.00) | 15  11.1  (6.88–42.7) | 15  138  (75.4–251) | 15  5307  (2151–13093) | 15  4273  (1863–9800) |
| **100 μg** | | | | | |
| **RBD ELISA**  **N**  **GMFR**  **(95% CI)** | 15  1.00  (1.00–1.00) | 15  205  (106–398) | 15  561  (265–1190) | 14  3528  (1638–7598) | 14  2343  (964–5697) |
| **S-2P ELISA**  **N**  **GMFR**  **(95% CI)** | 15  1.00  (1.00–1.00) | 15  657  (327–1318) | 15  831  (379–1820) | 14  6381  (2846–14310) | 14  6158  (3200–11850) |
| **ACOV2S**  **N**  **GMFR**  **(95% CI)** | 15  1.00  (1.00–1.00) | 15  128  (57.8–284) | 15  454  (312–660) | 14  19507  (15649–24317) | 14  13990  (11345–17252) |

N is the number of samples available for analysis.

GMFR, geometric mean fold rises.

**Supplementary Table 2. Summary of qualitative agreement measures between ACOV2S and neutralization assays (reference).**

|  | **nLUC_80_**  **(n=85)** | **PsVNA_50_**  **(n=144)** | **PRNT_80_***  **(n=57)** |
| --- | --- | --- | --- |
| **PPA** | 100 (92.5–100) | 100 (95.4–100) | 100 (ؘ87.2–100) |
| **NPA** | 78.9 (62.7–90.4) | 49.2 (36.6–61.9) | 100 (ؘ88.4–100) |
| **OPA** | 90.6 (82.3–95.8) | 77.1 (69.3–83.7) | 100 (ؘ93.7–100) |
| **PPV** | 85.5 (73.3–93.5) | 70.5 (61.2–78.8) | 100 (ؘ87.2–100) |
| **NPV** | 100 (88.4–100) | 100 (89.1–100) | 100 (ؘ88.4–100) |
| **Positive likelihood ratio (95% CI)** | 4.75 (2.57–8.79) | 1.97 (1.55–2.50) | Inf (–) |
| **Negative likelihood ratio (95% CI)** | 0 (–) | 0 (–) | 0 (–) |

Data shown as % (95% CI) unless otherwise stated.

* Only samples from Days 1 and 43 were available for comparison

**Supplementary Figure 1. Anti-SARS-CoV-2 (anti-N) assay-measured antibody levels following mRNA-1273 vaccination over time.** Line plots of the Elecsys Anti-SARS-CoV-2 assay cut-off indexes over time in samples of vaccinated trial participants, stratified by dose. Dotted grey vertical lines indicate time of vaccination, administered at Days 1 and 29. All results were below the reactivity cut-off (shown by red horizontal line).

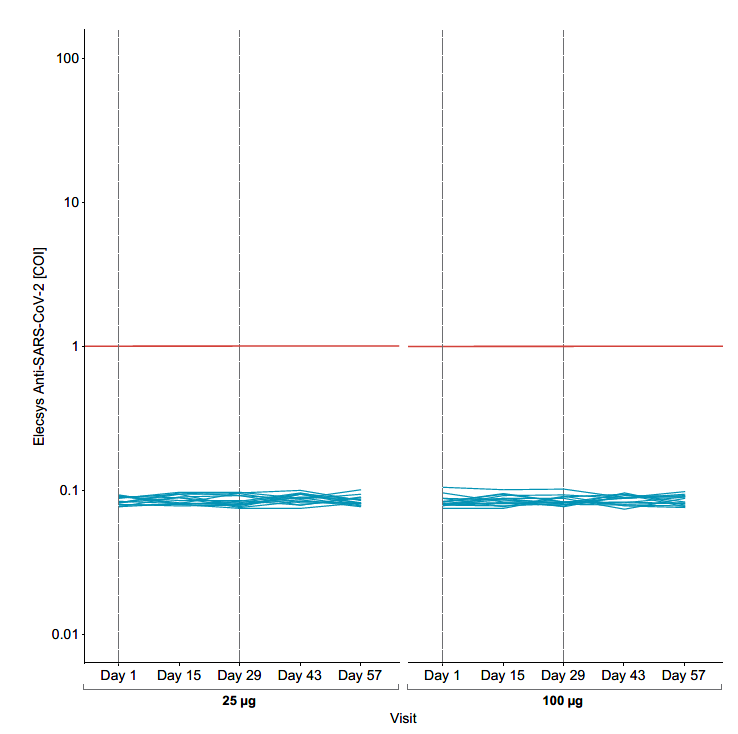
